# Adoption and Real-World Effectiveness of Adjunctive Azithromycin for Unscheduled Cesarean Delivery: A National Difference-in-Differences Analysis

**DOI:** 10.64898/2026.05.07.26352377

**Authors:** Taylor S. Freret, Ethan Litman, Timothy Wen, Jeanne-Marie Guise, Sarah E. Little, Mark A. Clapp

## Abstract

**Importance:** Cesarean delivery is the most common surgery in the US with more than 1 million performed each year; it is also the most significant risk factor for postpartum infection. The Cesarean Section Optimal Antibiotic Prophylaxis trial demonstrated that the addition of azithromycin at the time of cesarean birth performed in labor reduces postpartum infection.

**Objective:** To determine the real-world adoption and effect of this trial on clinical practice and postpartum infections among U.S. pregnant persons undergoing cesarean delivery in labor.

**Design:** Difference-in-differences analysis from 2013-2024.

**Setting:** Population-based, patient-level analysis using Epic Cosmos, a large longitudinal national electronic health record database of patients seen in health systems using Epic.

**Participants:** Pregnant individuals who received outpatient prenatal care in the system, who labored and gave birth to a liveborn singleton infant at 24-43 weeks of gestation were included. Exclusion criteria included unknown mode of delivery and intraamniotic infection.

**Exposures:** The treatment group included those delivered by cesarean and the control group included those who delivered vaginally. The pre-period was defined as 2013-2016, excluding a washout period from trial publication until December 31, 2016, and the post-period was defined from 2017-2024.

**Main Outcomes and Measures:** The primary outcomes were perioperative azithromycin administration and postpartum infection within 6 weeks of delivery.

**Results:** 1,663,341 participants were included in the final analysis. In the pre- and post-periods, azithromycin was administered in 0.01% and 0.04% of vaginal births and in 2.2% and 39.6% of cesarean births, respectively. In the pre- and post-periods, postpartum infection occurred in 2.0% and 2.7% of vaginal births and 9.2% and 8.0% of cesarean births. In the adjusted difference-in-difference analysis, the trial resulted in an absolute increase in azithromycin use by 37.6 percentage points (pp) (95% CI: 33.1 to 42.2 pp); postpartum infection decreased by 2.0 pp (95% CI: -2.5 to -1.4 pp), a relative decrease of 20%.

**Conclusions and Relevance:** Outside the clinical trial setting, this study provides evidence that azithromycin significantly reduces postpartum infection among pregnant persons undergoing a cesarean delivery in labor.

**Key Points:** *Question:* Did evidence from the Cesarean Section Optimal Antibiotic Prophylaxis (C/SOAP) trial change real-world clinical practice and decrease postpartum infections among U.S. pregnant persons who underwent a cesarean delivery in labor?

*Findings:* In this difference-in-differences analysis of 1.6 million births, azithromycin use increased 37.6 percentage points and postpartum infections decreased by 2.0 percentage points following the C/SOAP trial.

*Meaning:* Outside the clinical trial setting, this study provides evidence that azithromycin significantly reduces postpartum infection among individuals having a cesarean delivery in labor.

## Introduction

Postpartum infection is a major cause of maternal morbidity and mortality and associated with significant racial, ethnic, and socioeconomic disparities.^1-3^ Interventions to reduce infection are thus of significant public health importance. In the U.S., nearly one-third of all births occur by cesarean delivery, which remains the single most important risk factor for postpartum infection.^2,4^ Overall infection rates after cesarean delivery range from 10-20% and are nearly two-fold higher following cesarean deliveries performed in labor compared to prelabor cesarean deliveries.^2,5,6^ In 2024, more than 1.1 million individuals underwent cesarean delivery with nearly 30% occurring after a trial of labor, suggesting more than 60,000 pregnant individuals will develop a postpartum infection annually, even with the use of standard antibiotic prophylaxis.^7^

In 2016, the Cesarean Section Optimal Antibiotic Prophylaxis (C/SOAP) trial was published in the *New England Journal of Medicine*.^8^ This double-blind, pragmatic, randomized controlled trial of 2,000 women in the U.S. undergoing a cesarean delivery during labor compared rates of postpartum infection after randomization to intravenous azithromycin or placebo in addition to standard antibiotic prophylaxis. The trial demonstrated a 50% reduction in postpartum infection in the group that received azithromycin (6% vs 12%). Since 2016 when the trial was published, follow-up studies have supported the fetal safety and cost-effectiveness of azithromycin at the time of cesarean birth.^9,10^ However, there are no formal clinical guidelines recommending azithromycin for cesarean deliveries performed in labor; in its most recent guidance, the American College of Obstetricians and Gynecologists (ACOG) states that the intervention may be “considered.”^11^

In the absence of formal guidelines and given the challenges in implementing new evidence, it is not known to what extent azithromycin is used in practice following the C/SOAP trial. Moreover, there are often concerns about the external validity of trial findings and whether similar benefits will be seen when applied to a heterogeneous population that may not resemble the initial study.^12^ Thus, the objective of this study were to characterize the national uptake of azithromycin for unscheduled cesarean birth after publication of the C/SOAP trial and to determine whether there were corresponding decreases in postpartum infection rates.

## Methods

We conducted a difference-in-differences (DID) analysis using observational data in Epic Cosmos for births occurring between January 1, 2013, and December 31, 2024. Epic Cosmos is a dataset created in collaboration with a community of health systems using Epic representing more than 300 million patients from nearly 2,000 hospitals and 45,000 clinics as of March 2026.^13^ The dataset includes patients from all 50 states and Washington, D.C. The current count values for patients, hospitals, and clinics are available on cosmos.epic.com. The project was classified as non-human subjects research and was therefore exempt from institutional review board approval and informed consent. This study follows the STROBE reporting guidelines.^14^

Pregnant individuals who labored and gave birth to a liveborn singleton infant at 24 weeks 0 days to 43 weeks 6 days gestation between 2013 and 2024 were included. Labor was determined using the following variables in the Epic Cosmos data set: labor type (induction or spontaneous), labor start instant, first stage of labor length, second stage of labor start time or length, forceps or vacuum attempted, and vaginal delivery. Those with ≥ 240 minutes between rupture of membranes and delivery were also included, regardless of labor status, consistent with the eligibility criteria of the C/SOAP trial. Individuals were excluded if they did not have at least two outpatient visits in an obstetric, midwifery, or family medicine department during the pregnancy; this requirement was set to increase the likelihood of ascertaining postpartum infectious outcomes that may be managed in an office setting (e.g., wound infection). Consistent with the C/SOAP trial, we also excluded individuals with an encounter diagnosis for intraamniotic infection during the delivery encounter (ICD-10 O41.1). Furthermore, we excluded individuals whose mode of delivery (vaginal or cesarean) was unknown, who had rupture of membranes for more than 5 days (as a proxy for preterm prelabor rupture of membranes) or for an unknown duration, and those who did not have any perioperative antibiotics documented at the time of cesarean delivery (i.e., standard of care), as we could not be certain if this represented a documentation error or true omission of antibiotics.

The results of the C/SOAP trial were published in the *New England Journal of Medicine* on September 29, 2016. To complete the difference-in-differences analysis, we defined pre- and post-periods relative to publication date; the pre-period was January 2013 to September 28, 2016, and the post-period was January 2017 to December 2024. We also defined a washout period between September 29 and December 31, 2016, and excluded individuals delivering during this time to reduce the effects of early adoption. We extended the post-period through the end of 2024 to enable a contemporary analysis that also accounted for increasing adoption over time. As a sensitivity analysis, we performed additional analyses varying the duration of the post-period. As the trial findings were only relevant in individuals undergoing cesarean birth, we considered individuals who delivered by cesarean to be part of the treatment group and those who delivered by vaginal delivery as part of the control group.

The following patient characteristics were obtained from the EHR and compared between the periods in the treatment and control groups: maternal age, maternal race (categorized as American Indian or Alaskan Native, Asian, Black or African American, Native Hawaiian or Other Pacific Islander, White, other race, or multiracial), Hispanic ethnicity, parity, delivery body mass index (kg/m^2^, categorized as < 18.5, 18.5 – 24.9, 25-29.9, 30-39.9, or ≥ 40), prior cesarean birth, gestational age at delivery (categorized as 24-27, 28-31, 32-33, 34-36, 37-38, or ≥ 39 weeks), pregestational diabetes or gestational diabetes requiring medication (as diagnosed by at least two encounter ICD-10 codes, see Supplemental Table 1), rupture of membranes ≥ 18 hours, reaching the second stage, and insurance type. Patient demographics, including race, were examined to compare the patient characteristics in this study with those in the original trial.

The primary outcomes of interest were 1) use of perioperative azithromycin, and 2) a composite of postpartum infection or wound complication, modeled after the original C/SOAP trial. Intravenous azithromycin administered from 90 minutes before to 60 minutes after birth was identified in the EHR. Postpartum infections were identified by ICD-10 encounter diagnoses occurring within 42 days of the delivery admission (Supplemental Table 1). The composite outcome included any of the following: endometritis, sepsis, meningitis, pneumonia, other postpartum infection, pyelonephritis, wound infection, wound disruption, or urinary tract infection (UTI). As a sensitivity analysis, we also examined composite outcomes that excluded UTI and wound disruption, which were not included in the C/SOAP trial.

The difference-in-differences analysis used a patient-level linear regression model. Adjusted models controlled for age, parity, delivery BMI, diabetes, prior cesarean birth, gestational age, rupture of membranes ≥ 18 hours, based on *a priori* hypotheses and published risk factors for infection of labor or significant differences in the populations before and after the intervention.^15-18^ Individuals with missing covariate data were excluded from the adjusted analysis. Estimated marginal means were calculated by generating predicted values for each observation in the dataset at each treatment-period combination and then averaging across all observations. The parallel trends assumption was evaluated using two approaches: a visual inspection of pre-period trends and a test in which a linear model was fit to the pre-period data, and the interaction between time and treatment group tested for statistical significance. (Supplemental Table 2). Robust standard errors clustered by delivery site were used.

Data extraction was conducted utilizing Microsoft SQL Server Studio on July 26, 2025. Statistical analysis was conducted from July 2025 to March 2026. Analyses were performed in Rstudio 4.5.1. Standard mean differences (SMD) were calculated for univariate analyses, with < 0.10 considered evidence of balance. A 2-sided p-value < 0.05 was considered statistically significant.

## Results

There were 8,683,253 individuals with a recorded birth during the study period; 1,696,535 (19.5%) met criteria (Figure 1). There were 294,594 individuals who delivered in the pre-period; 33,094 in the washout period; and 1,368,847 in the post-period, for a total of 1,663,341 included in the final analysis. In the final cohort, 202,234 (12.2%) were delivered by cesarean. Births occurred at 1,951 unique delivery sites. Table 1 compares the characteristics of the study population in the pre- and post-periods, stratified by mode of delivery.

**Figure 1:**
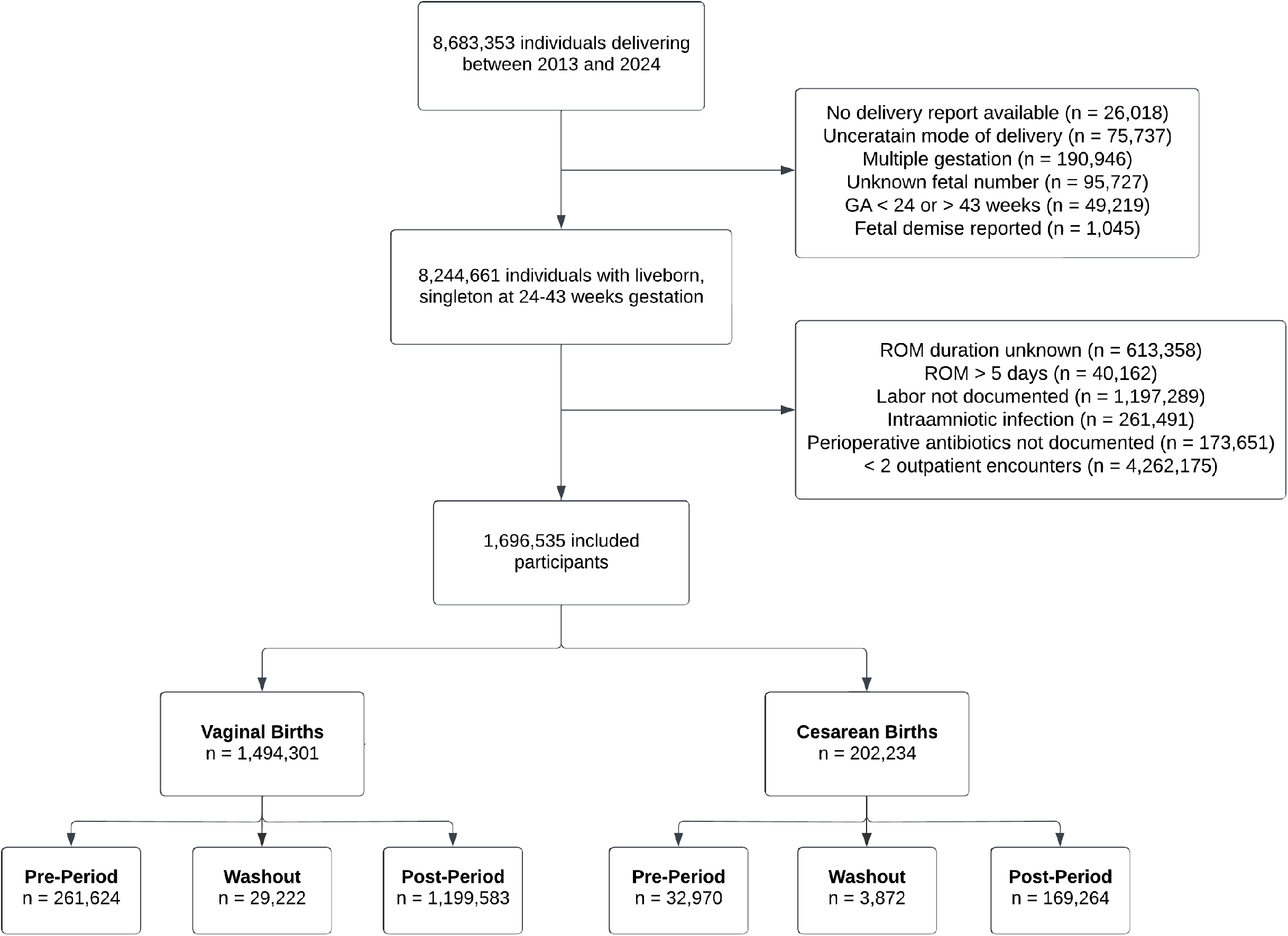
Study inclusion and exclusion criteria

**Table 1:** Baseline characteristics of the study population.

| Characteristic | Vaginal Birth |  |  | Cesarean Birth |  |  |
| --- | --- | --- | --- | --- | --- | --- |
|  | Pre-Period<br>(n = 261,624) | Post-Period<br>(n = 1,199,583) | SMD | Pre-Period<br>(n = 32,970) | Post-Period<br>(n = 169,264) | SMD |
| Age | 28.7 (5.7) | 29.3 (5.7) | 0.11 | 29.6 (6.0) | 30.2 (5.9) | 0.11 |
| Race |  |  |  |  |  |  |
| AIAN | 1,359 (0.5) | 4,816 (0.4) |  | 155 (0.5) | 705 (0.4) |  |
| Asian | 11,725 (4.5) | 51,464 (4.3) |  | 1,472 (4.5) | 7,362 (4.3) |  |
| Black or African American | 39,333 (15.0) | 183,389 (15.3) |  | 6,085 (18.5) | 30,496 (18.0) |  |
| NHOPI | 1,031 (0.4) | 5,756 (0.5) |  | 110 (0.3) | 756 (0.4) |  |
| Other Race | 19,221 (7.3) | 80,126 (6.7) |  | 2,247 (6.8) | 10,694 (6.3) |  |
| White | 152,964 (58.5) | 701,415 (58.5) |  | 17,507 (53.1) | 92,693 (54.8) |  |
| Multiracial | 32,391 (12.4) | 152,890 (12.7) |  | 4,952 (15.0) | 23,932 (14.1) |  |
| Missing | 3,600 (1.4) | 19,727 (1.6) | 0.04 | 442 (1.3) | 2,626 (1.6) | 0.05 |
| Hispanic ethnicity | 49,366 (18.9) | 215,688 (18.0) | 0.03 | 5,821 (17.7) | 29,022 (17.1) | 0.02 |
| Missing | 11,380 (4.3) | 55,144 (4.6) |  | 1,450 (4.4) | 8,074 (4.8) |  |
| Nulliparity | 94,828 (36.2) | 444,554 (37.1) | 0.02 | 18,544 (56.2) | 98,805 (58.4) | 0.04 |
| Delivery BMI (kg/m<sup>2</sup>) |  |  |  |  |  |  |
| < 18.5 | 65 (0.0) | 313 (0.0) |  | * | 20 (0.0) |  |
| 18.5-24.9 | 22,606 (8.6) | 84,735 (7.1) |  | 1,442 (4.4) | 5,700 (3.4) |  |
| 25-29.9 | 84,850 (32.4) | 350,943 (29.3) |  | 7,250 (22.0) | 32,234 (19.0) |  |
| 30-39.9 | 114,591 (43.8) | 557,325 (46.5) |  | 15,985 (48.5) | 82,598 (48.8) |  |
| ≥ 40 | 24,626 (9.4) | 145,413 (12.1) |  | 6,629 (20.1) | 41,209 (24.3) |  |
| Missing | 14,886 (5.7) | 60,854 (5.1) | 0.13 | 1,656 (5.0) | 7,503 (4.4) | 0.13 |
| Prior cesarean birth | 10,899 (4.2) | 53,160 (4.4) | 0.01 | 8,836 (26.8) | 43,045 (25.4) | 0.03 |
| ROM ≥ 18 hours | 16,625 (6.4) | 85,830 (7.2) | 0.03 | 5,416 (16.4) | 33,157 (19.6) | 0.08 |
| Pregestational diabetes | 3,645 (1.4) | 20,357 (1.7) | 0.03 | 1,436 (4.4) | 8,427 (5.0) | 0.03 |
| Gestational diabetes, A2 | 1,481 (0.6) | 39,573 (3.3) | 0.20 | 339 (1.0) | 8,801 (5.2) | 0.24 |
| Insurance Type |  |  |  |  |  |  |
| Public | 99,771 (38.1) | 483,600 (40.3) |  | 12,341 (37.4) | 63,564 (37.6) |  |
| Private | 100,017 (38.2) | 571,951 (47.7) |  | 13,608 (41.3) | 86,975 (51.4) |  |
| Other | 1,950 (0.7) | 14,777 (1.2) |  | 219 (0.7) | 1,928 (1.1) |  |
| Missing | 59,886 (22.9) | 129,255 (10.8) | 0.34 | 6,802 (20.6) | 16,797 (9.9) | 0.32 |
| Gestational Age (weeks) |  |  |  |  |  |  |
| 24-27 | 245 (0.1) | 902 (0.1) |  | 109 (0.3) | 517 (0.3) |  |
| 28-31 | 653 (0.2) | 2,531 (0.2) |  | 227 (0.7) | 1,022 (0.6) |  |
| 32-33 | 1,174 (0.4) | 5,256 (0.4) |  | 354 (1.1) | 1,784 (1.1) |  |
| 34-36 | 13,537 (5.2) | 63,448 (5.3) |  | 2,469 (7.5) | 14,382 (8.5) |  |
| 37-38 | 69,231 (26.5) | 358,093 (29.9) |  | 8,827 (26.8) | 51,805 (30.6) |  |
| ≥ 39 | 176,784 (67.6) | 769,353 (64.1) | 0.08 | 20,984 (63.6) | 99,754 (58.9) | 0.10 |

Data presented as mean (standard deviation) or n (%). The pre-period ran from Jan 1, 2013 to Sep 28, 2016, and the post-period from Jan 1, 2017 to Dec 31, 2024. AIAN: American Indian or Alaskan Native. NHOPI: Native Hawaiian or Other Pacific Islander. ROM: rupture of membranes. \*Counts < 11 are suppressed per Epic Cosmos guidelines.

The rate of perioperative azithromycin use increased significantly after 2016 among cesarean births (Figure 2). During the pre-period, 721/32,970 (2.2%) received azithromycin; in the post-period, 67,094/169,264 (39.6%) received azithromycin. By 2024, the rate was 59.7%. Use among vaginal births remained minimal throughout the study (pre-period: 15/261,624 [0.01%], post-period: 421/1,199,583 [0.04%]). In the unadjusted DID analysis, azithromycin use increased by 37.4 percentage points (95% CI: 32.9 to 41.9 pp) among cesarean births compared to vaginal births. Results were similar when adjusting for the covariates described above (adjusted DID estimate: 37.6 pp, 95% CI: 33.1 to 42.2; Table 2).

**Figure 2:**
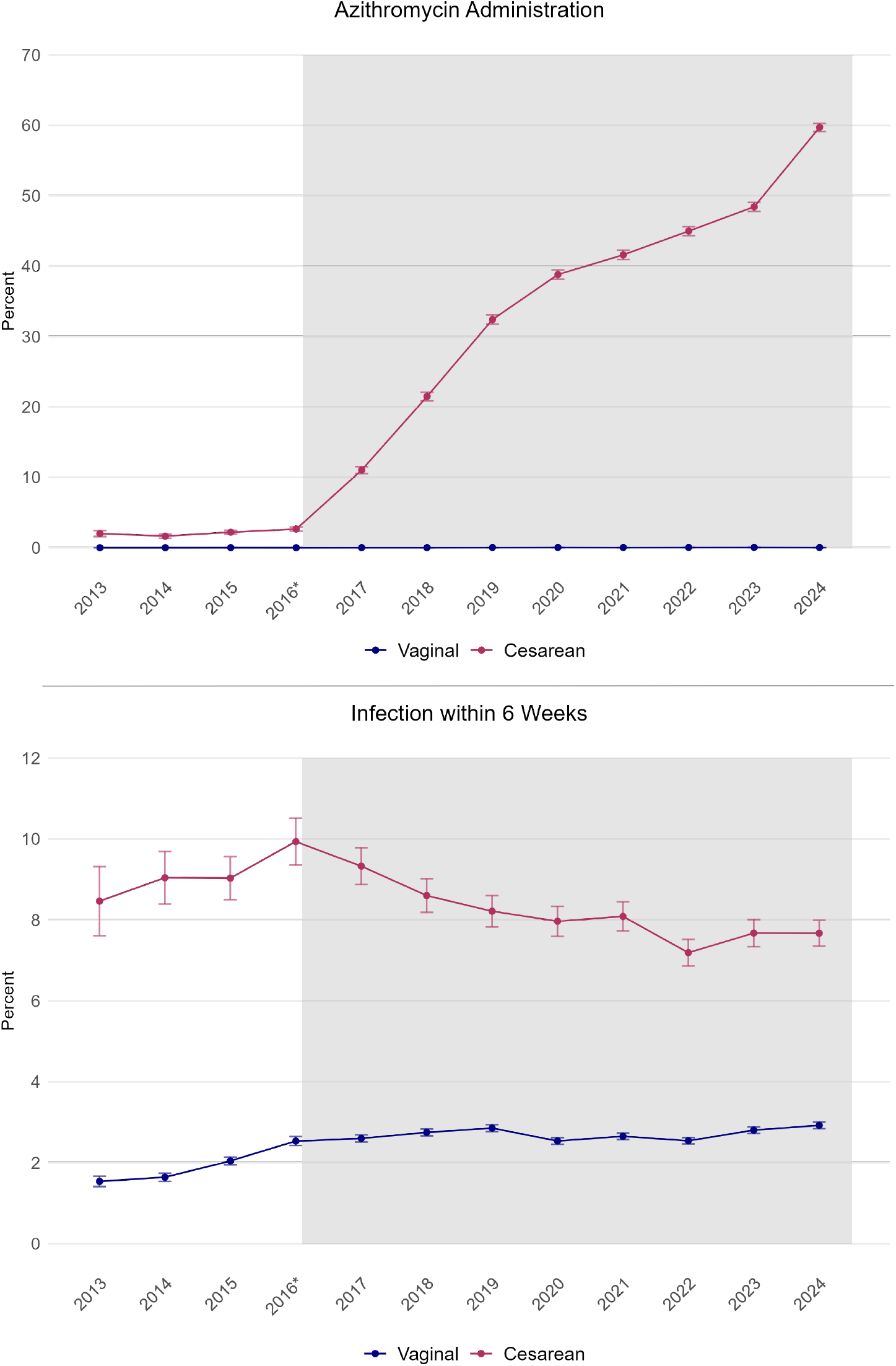
Trends in azithromycin administration and postpartum infection. Rates of azithromycin administration (top) and postpartum infection within 6 weeks (bottom) over time among individuals delivering vaginally or by cesarean after labor. Error bars indicate the 95% confidence interval. The gray shaded area indicates the post-period. *2016 excludes the washout period from Sep 29, 2016 to Dec 31, 2016.

**Table 2:** Unadjusted and adjusted DID estimates for azithromycin and postpartum infection among cesarean births in labor.

|  | **Pre-period**<br>**(n = 32,970)** | **Post-period**<br>**(n = 169,264)** | **Unadjusted DID**<br>**Estimate** | **Adjusted DID**<br>**Estimate** |
| --- | --- | --- | --- | --- |
| Azithromycin | 721 (2.2%) | 67,094 (39.6%) | 37.4 [32.9, 41.9] | 37.6 [33.1, 42.1] |
| Infection | 3,048 (9.6%) | 13,555 (8.1%) | -1.92 [-2.54, -1.31] | -2.00 [-2.61, -1.40] |
| Excluding UTI | 2,885 (9.1%) | 12,013 (7.2%) | -1.85 [-2.49, -1.20] | -1.91 [-2.54, -1.28] |
| Excluding UTI/WD | 2,307 (7.3%) | 9,414 (5.7%) | -1.62 [-2.23, -1.03] | -1.70 [-2.28, -1.11] |

Absolute counts and percent shown only among the cesarean birth group. Regression coefficients are multiplied by 100 to produce absolute rates. DID: difference-in-differences. UTI: urinary tract infection. WD: wound dehiscence.

Infection occurred after 2.0% of vaginal births in the pre-period and after 2.7% of vaginal births in the post-period. After cesarean delivery, infection occurred in 9.2% births in the pre-period and 8.0% in the post-period (Table 2). After the C/SOAP trial, postpartum infections after cesarean delivery decreased by -1.92 pp (95% CI: -2.54 to -1.31 pp). Findings were similar in the adjusted model (adjusted DID estimate: -2.00 pp, 95% CI: -2.61 to -1.40 pp) (Table 2). Compared to the expected rate of 10.0%, the DID estimate of -2.00 represents a 20% relative decrease. Findings were similar for the outcomes excluding wound disruption and UTI, or UTI alone (Table 2). When varying the length of the post-periods, the absolute value of the DID estimate progressively increased, consistent with increasing adoption over time (Supplemental Table 3).

Rates of the individual infection components in the pre- and post-periods are presented in Table 3, stratified by mode of delivery. In the post-period among cesarean deliveries, there were significant reductions across a variety of infections, including surgical site infections like wound infection or disruption. However, there was also a reduction in more significant infections: rates of sepsis decreased by 50% and endometritis by 25%. Although rates of UTI were higher in the post-period among cesarean births, this trend was also seen among vaginal births.

**Table 3:** Absolute rates of infections by mode of delivery in the pre- and post-period.

|  | Vaginal |  |  | Cesarean |  |  |
| --- | --- | --- | --- | --- | --- | --- |
|  | Pre-period<br>(n = 261,624) | Post-period<br>(n = 1,199,583) | P | Pre-period<br>(n = 32,970) | Post-period<br>(n = 169,264) | P |
| **Any Infection** | 5,311 (2.0) | 32,581 (2.7) | < 0.001 | 3,048 (9.2) | 13,555 (8.0) | < 0.001 |
| **Sepsis** | 1,554 (0.6) | 5,438 (0.5) | < 0.001 | 608 (1.8) | 1,467 (0.9) | < 0.001 |
| **Endometritis** | 1,494 (0.6) | 7,363 (0.6) | 0.01 | 655 (2.0) | 2,541 (1.5) | < 0.001 |
| **Wound infection** | 329 (0.1) | 1,576 (0.1) | 0.46 | 1,214 (3.7) | 5,277 (3.1) | < 0.001 |
| **Wound disruption** | 241 (0.1) | 1,105 (0.1) | 1.00 | 901 (2.7) | 4,101 (2.4) | 0.001 |
| **Other PPI** | 1,667 (0.6) | 9,648 (0.8) | < 0.001 | 490 (1.5) | 1,954 (1.2) | < 0.001 |
| **UTI** | 1,330 (0.5) | 13,138 (1.1) | < 0.001 | 222 (0.7) | 2,089 (1.2) | < 0.001 |
| **Pyelonephritis** | 319 (0.1) | 2,083 (0.2) | < 0.001 | 1,214 (3.7) | 251 (3.1) | 0.08 |
| **Meningitis** | 16 (0.0) | 79 (0.0) | 0.78 | * | 13 (0.0) | 0.49 |
| **Pneumonia** | 288 (0.1) | 1,316 (0.1) | 0.96 | 108 (0.3) | 445 (0.3) | 0.06 |

Data presented as n (%). P-value < 0.05 considered statistically significant. PPI: postpartum infection. UTI: urinary tract infection. \*Counts < 11 are suppressed per Epic Cosmos guidelines.

## Discussion

In summary, the C/SOAP trial increased the use of azithromycin for cesarean birth after labor by 37.6 percentage points and decreased infection by 2.04 percentage points, representing a 20% reduction in postpartum infections. Findings were robust to covariate adjustment and varying post-period specification lengths, demonstrating the robustness of this quasi-experimental approach. Although randomized clinical trials are the gold standard for determining the effectiveness of an intervention, their translation into clinical practice can be limited by issues of generalizability outside highly controlled environments.^19,20^ Our study suggests that at a population level, perioperative azithromycin use at the time of cesarean delivery in labor decreases postpartum infection, findings that enhance the external validity of the C/SOAP trial findings.

Although our study supports the real-world effects of the C/SOAP trial, there are some notable differences in the populations. For example, the postpartum infection rate in the C/SOAP trial was 12% in the control group and 6% in those receiving azithromycin. In contrast, we observed lower postpartum infection rates in the pre-period (9.2%). This is likely in part due to the use of ICD codes to identify postpartum infection, as well as differences in the study populations. For example, our study population included a much higher rate of individuals with private insurance (approximately 45% vs 30% in the C/SOAP trial); prior studies have suggested that postpartum infection rates are lower in a privately insured population.^21,22^

We also observed a smaller decrease in postpartum infection seen in our study following azithromycin (20% vs 49% in the C/SOAP trial). The lower reduction can be attributed in part to the fact that less than 50% of individuals in the post-period received azithromycin, compared with 100% in the C/SOAP azithromycin group. Therefore, we would not expect the average effect in the post-period to mirror that of the C/SOAP intervention group, given that practice adoption is increasing over time. Additionally, the use of antibiotics in the real world does not exactly recreate their use in a highly controlled clinical trial. Given that more than 300,000 individuals have a cesarean delivery in labor annually, a 20% reduction still represents a meaningful improvement in postpartum outcomes, potentially preventing 6,000 infections each year.^7^

There are several strengths to our study. This study leverages a large, contemporary, national electronic health record database (Epic Cosmos) that includes births from all 50 states across academic and community hospitals in addition to detailed birth and prescription data, enhancing the generalizability of our findings. The use of quasi-experimental DID methodology lends robustness to the findings. In particular, the inclusion of a control group (vaginal births) unaffected by the dissemination of the C/SOAP trial tempers the effects of differential outcome assessment over time (e.g., the ICD-10 transition that occurred in October 2015). We hypothesize that the increasing rates of postpartum infection seen from 2013-2016 were partly due to the increased (and eventually required) use of ICD-10 codes, which appears supported by stable rates in the vaginal delivery group after 2016. Reassuringly, any issues related to the ICD transition would have affected both groups equally.

There are also several limitations to our study. Although quasi-experimental DID analysis can be used to assess causality, the conclusions are valid only when no other interventions affect the measured outcomes. To our knowledge, no other factors would have affected azithromycin use in the post-period; however, we cannot exclude the possibility that other interventions (e.g., vaginal prep) may be changing in the post-period, thereby reducing the incidence of postpartum infection. However, the clear demonstration of azithromycin adoption and a corresponding change in infection rates following the C/SOAP trial publication supports the assertion that a significant portion of the reduction can be attributed to increased azithromycin use. Epic Cosmos does not provide access to all clinical information that is captured during a delivery encounter. In particular, the indication for cesarean delivery was not available at the time of the study, nor was the scheduled or unscheduled nature of the delivery. We used proxy measures to identify individuals who underwent unscheduled cesarean birth (e.g., during labor). Another limitation of our study is the reliance on ICD-10 codes to identify infectious outcomes. We mapped infectious complications from the C/SOAP trial to corresponding ICD-10 codes and increased the likelihood of capturing outpatient postpartum diagnoses by excluding individuals without antenatal outpatient encounters in Cosmos. Assuming the codes were not differentially used after cesarean and vaginal births, the effects reported correspond to meaningful changes in the infectious outcomes rather than to the study’s use of ICD-10 codes.

## Conclusion

In conclusion, this quasi-experimental DID analysis using the Epic Cosmos system demonstrated that the publication of the C/SOAP trial significantly increased the use of azithromycin for cesarean birth after labor and reduced postpartum infection. Further research examining effects in subpopulations that may particularly benefit from azithromycin (such as individuals with obesity or preexisting diabetes) or in groups excluded from the original trial (multiple gestation pregnancies, pregnancies complicated by intraamniotic infection) would help further elucidate the real-world effects of the C/SOAP trial and of azithromycin.

## Data Availability

Access to Epic Cosmos data is restricted by Epic and therefore data will not be shared.

## Acknowledgments

TSF had full access to all the data in the study and takes responsibility for the integrity of the data and the accuracy of the data analysis.

There was no funding provided for this study.

This work was presented as an oral presentation at the Society for Maternal Fetal Medicine Annual Meeting on February 12, 2026, in Las Vegas, NV.

Disclosures: Dr. Clapp and Dr. Wen serve as medical advisory board members with private equity in Delfina Health, outside the submitted work. Dr. Clapp also reports grants from the NHLBI and AHRQ, outside the submitted work. Dr. Clapp receives payment for serving on a data safety monitoring board from Novocuff, Inc., and also receives a stipend from the American College of Obstetricians and Gynecologists for editorial services. Dr. Wen receives a stipend from the American Journal of Obstetrics and Gynecology MFM for editorial services.

## Notes

### Funding Statement

This study did not receive any funding.

### Author Declarations

The Beth Israel Deaconess Medical Center institutional review board waived ethical approval as the study is not considered human subjects research. (Data source is Epic Cosmos, where all patient information is de-identified and therefore is no identifiable patient information.)

## References

1. Pregnancy-Related Deaths: Data from Maternal Mortality Review Committees. CDC. Accessed March 10, 2026. https://www.cdc.gov/maternal-mortality/php/data-research/mmrc/?CDC_AAref_Val=https%3A%2F%2Fwww.cdc.gov%2Fmaternal-mortality%2Fphp%2Fdata-research%2Findex.html&cove-tab=1#:~:text=Underlying%20causes%20of%20pregnancy%2Drelated%20deaths%2C%20overall%20and,death%20for%20the%20selected%20race%20or%20ethnicity

2. Tita ATN, Rouse DJ, Blackwell S, Saade GR, Spong CY, Andrews WW. Emerging concepts in antibiotic prophylaxis for cesarean delivery: a systematic review. Obstetrics and gynecology. Mar 2009;113(3):675–682. doi:10.1097/AOG.0b013e318197c3b6

3. Williams A, Little SE, Bryant AS, Smith NA. Mode of Delivery and Unplanned Cesarean: Differences in Rates and Indication by Race, Ethnicity, and Sociodemographic Characteristics. Am J Perinatol. May 2024;41(7):834–841. doi:10.1055/a-1785-8843

4. Osterman MJK, Hamilton BE, Martin JA, Driscoll AK, Valenzuela CP. Births: Final Data for 2023. Natl Vital Stat Rep. Mar 18 2025;(1):1. doi:10.15620/cdc/175204

5. Duff P. Infection after cesarean delivery: diagnosis, pathophysiology, management, and prevention. American journal of obstetrics and gynecology. Jan 2026;233(6S):S464–S482. doi:10.1016/j.ajog.2025.08.007

6. Lamont RF, Sobel JD, Kusanovic JP, et al. Current debate on the use of antibiotic prophylaxis for caesarean section. BJOG. Jan 2011;118(2):193–201. doi:10.1111/j.1471-0528.2010.02729.x

7. Centers for Disease Control and Prevention, National Center for Health Statistics. National Vital Statistics System, Natality on CDC WONDER Online Database. Accessed April 22, 2026. http://wonder.cdc.gov/natality-expanded-current.html

8. Tita AT, Szychowski JM, Boggess K, et al. Adjunctive Azithromycin Prophylaxis for Cesarean Delivery. N Engl J Med. Sep 29 2016;375(13):1231–41. doi:10.1056/NEJMoa1602044

9. Martin JK, Longo SA, Jauk VR, et al. Neonatal outcomes in term and preterm infants following adjunctive azithromycin antibiotic prophylaxis for non-elective cesarean delivery. J Matern Fetal Neonatal Med. Dec 2024;37(1):2367082. doi:10.1080/14767058.2024.2367082

10. Harper LM, Kilgore M, Szychowski JM, Andrews WW, Tita ATN. Economic Evaluation of Adjunctive Azithromycin Prophylaxis for Cesarean Delivery. Obstetrics and gynecology. Aug 2017;130(2):328–334. doi:10.1097/AOG.0000000000002129

11. ACOG Practice Bulletin No. 199 Summary: Use of Prophylactic Antibiotics in Labor and Delivery. Obstetrics and gynecology. Sep 2018;132(3):798–800. doi:10.1097/AOG.0000000000002834

12. Petersen MK, Andersen KV, Andersen NT, Soballe K. “To whom do the results of this trial apply?” External validity of a randomized controlled trial involving 130 patients scheduled for primary total hip replacement. Acta Orthop. Feb 2007;78(1):12–8. doi:10.1080/17453670610013367

13. Epic Cosmos. Epic. Accessed March 15, 2026. https://cosmos.epic.com/

14. von Elm E, Altman DG, Egger M, et al. The Strengthening the Reporting of Observational Studies in Epidemiology (STROBE) statement: guidelines for reporting observational studies. Lancet. Oct 20 2007;370(9596):1453–7. doi:10.1016/S0140-6736(07)61602-X

15. Hayasaka M, Landy HJ, Forbes L, et al. Surgical site infections after cesarean delivery. American journal of obstetrics and gynecology. Jan 2026;233(6S):S524–S540. doi:10.1016/j.ajog.2025.08.008

16. Leth RA, Uldbjerg N, Norgaard M, Moller JK, Thomsen RW. Obesity, diabetes, and the risk of infections diagnosed in hospital and post-discharge infections after cesarean section: a prospective cohort study. Acta Obstet Gynecol Scand. May 2011;90(5):501–9. doi:10.1111/j.1600-0412.2011.01090.x

17. Haidar ZA, Nasab SH, Moussa HN, Sibai BM, Blackwell SC. Caesarean Delivery Surgical Site Infection: What are Expected Rates and Potentially Modifiable Risk Factors? Journal of obstetrics and gynaecology Canada : JOGC = Journal d’obstetrique et gynecologie du Canada : JOGC. Jun 2018;40(6):684–689. doi:10.1016/j.jogc.2017.09.020

18. Reddy UM, Rice MM, Grobman WA, et al. Serious maternal complications after early preterm delivery (24-33 weeks’ gestation). American journal of obstetrics and gynecology. Oct 2015;213(4):538 e1–9. doi:10.1016/j.ajog.2015.06.064

19. Fernainy P, Cohen AA, Murray E, Losina E, Lamontagne F, Sourial N. Rethinking the pros and cons of randomized controlled trials and observational studies in the era of big data and advanced methods: a panel discussion. BMC Proc. Jan 18 2024;18(Suppl 2):1. doi:10.1186/s12919-023-00285-8

20. Gerstman BB. There is no single gold standard study design (RCTs are not the gold standard). Expert Opin Drug Saf. Apr 2023;22(4):267–270. doi:10.1080/14740338.2023.2203488

21. Taylor YJ, Liu TL, Howell EA. Insurance Differences in Preventive Care Use and Adverse Birth Outcomes Among Pregnant Women in a Medicaid Nonexpansion State: A Retrospective Cohort Study. J Womens Health (Larchmt). Jan 2020;29(1):29–37. doi:10.1089/jwh.2019.7658

22. Yi SH, Perkins KM, Kazakova SV, et al. Surgical site infection risk following cesarean deliveries covered by Medicaid or private insurance. Infect Control Hosp Epidemiol. Jun 2019;40(6):639–648. doi:10.1017/ice.2019.66

