## Supplemental Tables for "Adoption and Real-World Effectiveness of Adjunctive Azithromycin for Unscheduled Cesarean Delivery: A National Difference-in-Differences Analysis"

Supplemental Table 1:

| **Infection Type** | **ICD Codes** |
| --- | --- |
| Endometritis | O86.12 |
| Meningitis | G00, G01, G02, G03, G06, G07 |
| Pneumonia | J13, J14, J15, J16, J17, J18 |
| Other Postpartum Infection | O86.1 except O86.12, O86.4, O86.8 |
| Pyelonephritis | N10, O86.21 |
| Wound Infection | O86.0 except O86.04, T81.4 except T81.44 |
| Sepsis | A40, A41, R65.2, O85, O86.04, T81.44 |
| Wound Disruption | O90.0, T81.3 |
| Urinary tract infection | O86.2 except O86.21, N30.0 |
| Pregestational Diabetes | O24.0, O24.1, E10, E11 |
| Gestational Diabetes, A2 | O24.414, O24.415, O24.424, O24.425, O24.434, O24.435 |

All subcodes included unless specifically noted.

Supplemental Table 2: Parallel trends testing

| **Model** | **Slope Difference Estimate** | **p-value** |
| --- | --- | --- |
| Azithromycin | 0.3 [-0.3, 0.9] | 0.30 |
| Any Infection | 0.1 [-0.3, 0.5] | 0.66 |
| Infection without UTI | 0.2 [-0.1, 0.5] | 0.24 |
| Infection without UTI or WD | 0.0 [-0.4, 0.4] | 0.94 |

Slope difference provides the estimated difference in the pre-period trends among cesarean and vaginal births prior to the intervention. None of the estimates are statistically significant; if they were, this would suggest that the parallel trends assumption was violated.

Supplemental Table 3: Unadjusted DID estimates obtained with varying post-period duration

|  | **Azithromycin** | **Any Infection** |
| --- | --- | --- |
| **Primary Analysis** | 37.4 [32.9, 41.9] | -1.9 [-2.5, -1.3] |
| **Sensitivity Analysis** |  |  |
| Post-period length = 1 year | 8.8 [5.7, 11.9] | -0.5 [-1.2, -0.2] |
| Post-period length = 2 years | 14.3 [10.8, 17.8] | -0.9 [-1.6, -0.3] |
| Post-period length = 3 years | 20.2 [16.3, 24.0] | -1.3 [-1.9, -0.7] |
| Post-period length = 4 years | 24.8 [20.6, 28.9] | -1.4 [-2.0, -0.8] |
| Post-period length = 5 years | 28.2 [23.7, 32.7] | -1.5 [-2.1, -0.9] |
| Post-period length = 6 years | 31.1 [26.4, 35.8] | -1.7 [-2.3, -1.1] |
| Post-period length = 7 years | 33.7 [28.9, 38.4] | -1.8 [-2.4, -1.2] |

Sensitivity analysis varying the length of the post-period (i.e., number of years after the publication of the C/SOAP trial).
